# Prevalence of Chikungunya, Dengue, and West Nile arboviruses in Iran based on enzyme-linked immunosorbent assay (ELISA): A systematic review and meta-analysis

**DOI:** 10.1101/2024.09.12.24313525

**Authors:** Ebrahim Abbasi, Mohammad Djaefar Moemenbellah-Fard

## Abstract

**Introduction:** Arboviruses, including Chikungunya (CHIKV), Dengue (DENV), and West Nile (WNV) viruses, are significant viral threats that affect numerous people globally each year. This report explore the prevalence of these viruses in Iran through a systematic review and meta-analysis.

**Methods:** The present survey was performed with a systematic review and meta-analysis method on the seroprevalence of WNV, CHIKV, and DENV using the ELISA test. Based on this, by search in Web of Science, PubMed, Scopus, Cochrane Library, Science Direct, and Google Scholar scientific databases, all relevant published papers were sorted out and reviewed. Power ratification of data was carried out with a random effects model in meta-analysis, meta-regression, *I*^*2*^ index, and Egger test.

**Results:** Twelve published papers between 2000 and 2024 were embodied in this meta-analysis report. The seroprevalence of positive ELISA test for WNV in Iran was estimated at 12.9% (CI=95%: 7.4-18.4), and for CHIKV at 6.2% (CI=95%: 0.6-11.8). Regarding DENV, only two studies were conducted with a zero prevalence in one study, and a seroprevalence of 5.6% in another study.

**Conclusion:** According to these data, WNV, CHIKV, and DENV fevers have been detected in Iran using ELISA test. Considering the seropositivity of WNV and CHIKV, and the finding of these viruses from several provinces, it could be for granted that these two viruses are ubiquitous and DENV fever is sporadic in Iran.

**Author Summary:** Arboviruses, including Chikungunya (CHIKV), Dengue (DENV), and West Nile (WNV), pose significant health threats globally, with increasing infections reported, including in Iran. This report conducts a systematic review and meta-analysis of the seroprevalence of these viruses in Iran through ELISA testing. A total of twelve studies published from 2000 to 2024 were analyzed, revealing a seroprevalence of 12.9% for WNV and 6.2% for CHIKV, while DENV showed a mixed prevalence of 0% in one study and 5.6% in another. The results suggest that WNV and CHIKV are widespread in Iran, whereas DENV appears to be sporadic.

## Introduction

West Nile virus (WNV) is a single strand positive sense RNA virus in the Flaviviridae family, which is mainly transmitted by *Culex* and *Aedes* mosquitoes. *Culex pipiens, Culex tarsalis*, and *Aedes vexans* are the main vectors of WNV ^1 2^. In Iran, *Cx. pipiens* and *Ae. caspius* are vectors of WNV. Trans-ovarian transmission of WNV among mosquito vectors is also possible, which renders them reservoirs of this pathogen. WNV circulates between insects, birds, and mammals (as the dead-end host), and their life is maintained in a cycle of “amplification” ^2^. The virus cannot transfer between mammals, especially humans to humans, and can only be transmitted to another person through organ transplants or blood transfusions, and placental/ milk transmission. Accidental human infection is often (≈80%) asymptomatic, in 20% of cases with mild flu-like symptoms, and <1% can cause WNV neurotropic disease (WNND, West Nile Neuro-invasive Disease), which usually occurs in elderly and immunocompromised individuals, that can lead to death ^3-6^.

Migratory birds play important roles in the transmission and spread of WNV in the world, so today, this virus is prevalent in most parts of the world, including Europe and Asia. Since no effective and safe vaccine is available to deal with this virus ^7^, organizations and departments related to health have suggested that WNV monitoring plans be implemented in the form of collecting virologic, entomologic, veterinary, and epidemiologic data to monitor the spread of WNV, and identify cases of human infection so that prevention, control, and adoption of appropriate treatment protocols to deal with this virus could be undertaken ^8-10^.

Dengue virus (DENV) is also a single strand positive sense RNA virus in the Flaviviridae family, and the causative agent of the newly-emergent dengue fever which has four serotypes 1, 2, 3, and 4 ^11^. The primary and secondary vectors of DENV are *Aedes aegypti* and *Aedes albopictus* mosquitoes ^12^. Trans-ovarian transmission of DENV among vectors is possible. The life of all four DENV serotypes is maintained in two sylvatic and urban cycles. The Asian tiger mosquito, *Ae. albopictus*, serves as a “bridge” vector between these two cycles. Both these diurnally-active anthropophilic *Aedes* vectors are increasingly reported from SE Iran. In the sylvatic cycle, the virus is transmitted from non-human mammals to mosquitoes, and then to mammals, such as the monkey-*Aedes*-monkey cycle, which is ecologically and evolutionarily distinct from the human transmission cycle ^13^.

More than 400 million infections, and >20,000 deaths occur annually due to DENV worldwide ^11 14^. This disease often exists as a latent or asymptomatic infection, and in endemic/ epidemic forms in different regions of the world ^11^. Low-to-medium viremia in these persons could be infectious to mosquitoes. The continents of Asia, America, Africa, and Australia witness DENV epidemics every year ^15^. Clinical manifestations of dengue infection range from mild fever to severe dengue hemorrhagic fever (DHF). Infection with one serotype of DENV can provide lifelong immunity to the same serotype, while secondary infection with other serotypes levitates the risk of infection to severe DENV. No effective vaccine is available for this virus ^16^.

Chikungunya virus (CHIKV) is a single strand positive sense RNA virus of the alphavirus genus in Togaviridae family. Alphaviruses can cause inflammatory musculoskeletal diseases with debilitating symptoms such as arthritis, arthralgia, and myalgia in humans ^17^. CHIKV is transmitted by *Aedes* mosquitoes, especially *furcifer, africanus, aegypti, albopictus*, and *Stegomyia* species ^18 19^. This virus has three genotypes, namely West Africa (WA), East/Central/Southern Africa (ECSA), and Asian. All three genotypes are distributed worldwide, but ECSA and Asian genotypes are more common ^19-21^. The virus is maintained in a rural enzootic transmission cycle or the sylvatic cycle between *Aedes* mosquitoes and animal reservoirs. However, today the virus has adapted to urban cycles, and no longer requires the presence of non-human primates or the sylvatic cycle for their maintenance ^22^.

Unlike DENV, CHIKV is not life threatening. Mortality caused by this virus is low, but it can cause severe complications, and affect people’s quality of life. In most cases, CHIKV infection presents with a sudden onset of fever accompanied by joint pain. In minor cases, it leads to polyarthralgia and debilitating arthritis, rashes, myalgia, and headache ^23^. Asymptomatic infection is rare and occurs in 3-28% of cases in epidemics ^24 25^. The infection is often self-limiting, and the patient eventually recovers, but some patients develop persistent joint pain that may persist for months or years after the acute stage of the disease ^26 27^. Approximately 30–40% of infected individuals experience some long-term complications ^28^. So far, no vaccine is available to prevent this virus ^18^.

ELISA (Enzyme-linked immunosorbent assay) test is a method of serological examination of viruses in the world. Due to its high sensitivity, and low false positive rate of about 20%, this test is recommended as a serological test for the initial screening of viral diseases. The presence of DENV IgG antibodies in people indicates that they have been infected with the virus in the past or present, which is usually investigated in the ELISA test ^29^.

Due to the lack of an effective vaccine to prevent viral infections of arboviruses and the existence of complications and death caused by these viruses, awareness of the prevalence, distribution, and detection methods of these viruses to adopt control and prevention programs is essential. The named viral diseases have been known in Iran in the past years. However, their prevalence is different in different regions of Iran. Accordingly, this study was conducted to determine the positive rate of ELISA test for WNV, DENV, and CHIKV by a systematic review and meta-analysis method in Iran, to achieve comprehensive results regarding the spread of these viruses.

## Materials and Methods

### Study Protocol

This study was conducted by the method of systematic review and meta-analysis in the field of ELISA tests for the three arboviruses of WNV, CHIKV, and DENV based on the guidelines of PRISMA (Preferred Reporting Items for Systematic Reviews and Meta-Analyses) ^30^.

### Search Strategy

In the initial search, all English-language articles published from the beginning of 2000 to the end of May 2024 were extracted by searching the Web of Science, PubMed, Scopus, Cochrane Library, Science Direct, and Google Scholar databases. Search for articles using the keywords Arbovirus, West Nile Virus, West Nile Virus Infection, WNV, West Nile Virus IgG, IgG anti-WNV, Dengue Fever, Dengue virus, Dengue virus infection, Chikungunya virus, Enzyme-linked immunosorbent assay, ELISA, Serology, Seroprevalence, Seropositivity, Iran, Iranian in the title, abstract and keywords were done in singular and compound form using “AND” and “OR” operators.

### Inclusion and Exclusion Criteria

All the English-language articles published in Iran were in the field of arboviruses: WNV, CHIKV, and DENV. The serological examination of the virus was done using the ELISA test, the positive rate of ELISA test was reported with good quality. These were entered into the study. Articles that did not meet the inclusion criteria or were conducted using meta-analysis, review, case report, or case series methods were excluded from the study.

### Quality Assessment

The quality assessment of the articles was done based on 22 parts of the STROBE (Strengthening the Reporting of Observational Studies in Epidemiology) checklist, which examined compliance with the principles of writing and implementation in the title, the method of reporting findings, limitations, and conclusions. Each part of this checklist is given a score based on its importance, and the maximum possible score is 33 ^31^.

### Screening and Data Extraction

The articles were screened by two researchers independently by examining the title and abstract of the articles and considering the inclusion and exclusion criteria. Finally, the full text of the articles was investigated by two researchers independently, and if the articles were rejected by two people, the reason was mentioned, and in case of disagreement between them, the article was refereed by a third person. Data extraction was done using a pre-prepared checklist that included study location, study time, sample size, type of serological test, number of positive ELISA tests, and type of virus under investigation.

### Study Selection

In the initial search and considering the inclusion criteria, 794 articles were extracted. Then, using the Endnote software, the duplication of sources was investigated, and 259 articles were excluded from the study due to duplication. Then, by investigating the titles and summaries of the articles, 486 articles were excluded from the study due to their irrelevance. After reviewing the full text of the articles, 37 articles were excluded due to the lack of investigation on the prevalence of the ELISA test and the unknown population. Finally, 12 articles entered the meta-analysis process (Figure 1).

**Figure 1.**
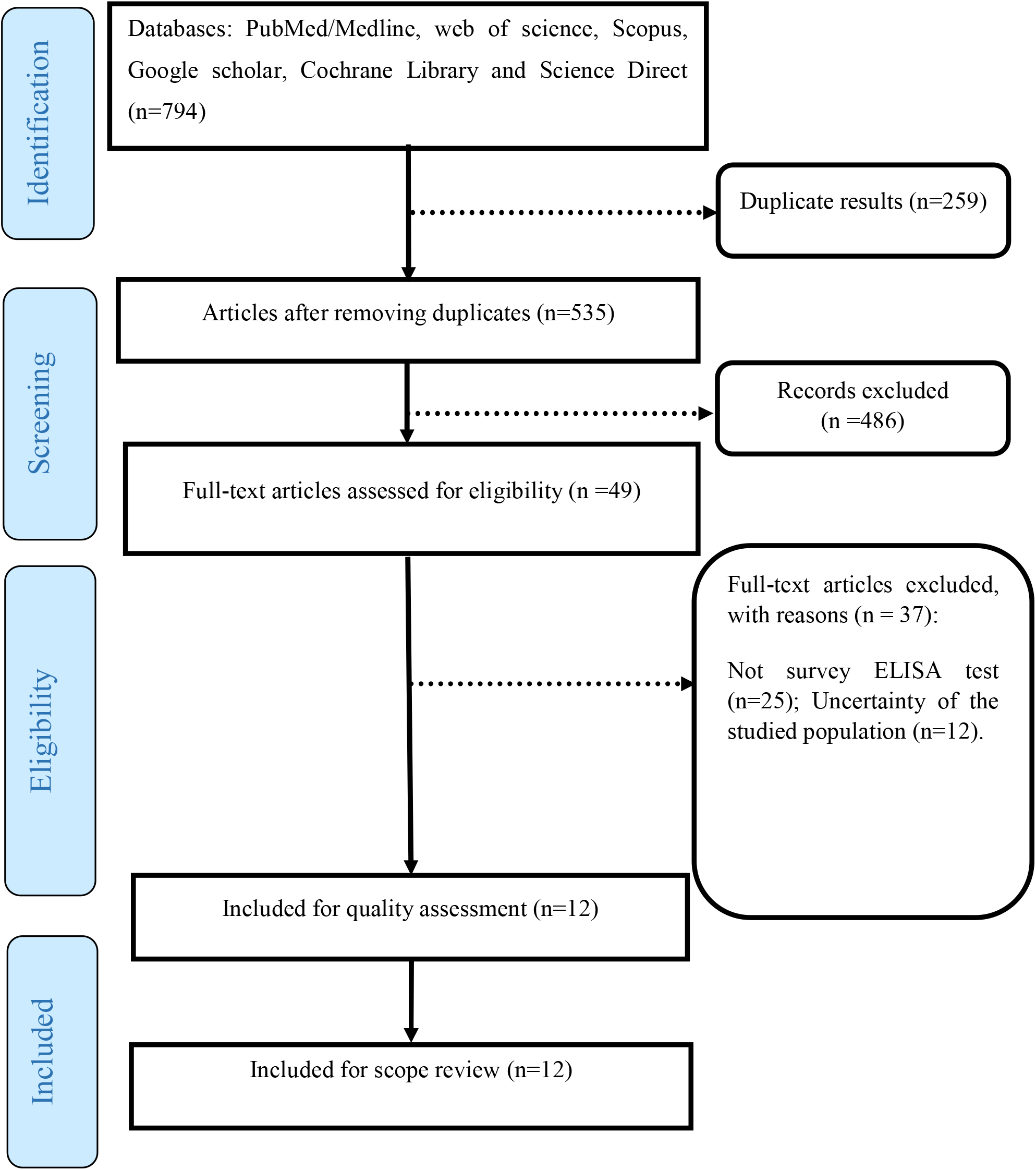
The review process based on PRISMA flow chart

### Statistical Analysis

In order to combine the results in heterogeneous studies, random-effects model was used, and in the homogeneous studies, the fixed-effects model in meta-analysis was deployed. To investigate the heterogeneity of the data, *I*^*2*^ and Cochrane Q tests were used. Publication bias was controlled by Egger test, and funnel plot and data analysis were performed using STATA ver. 17.0 software.

## Results

Overall, 12 articles that were conducted in Iran between 2010 and 2023 were included in this meta-analysis process. Nine articles on WNV ^29 32-39^, three articles on CHIKV ^38 40 41^, and two articles on DENV ^29 42^ were scrutinized. The characteristics of the articles included in the meta-analysis are presented in Table 1.

**Table 1.**
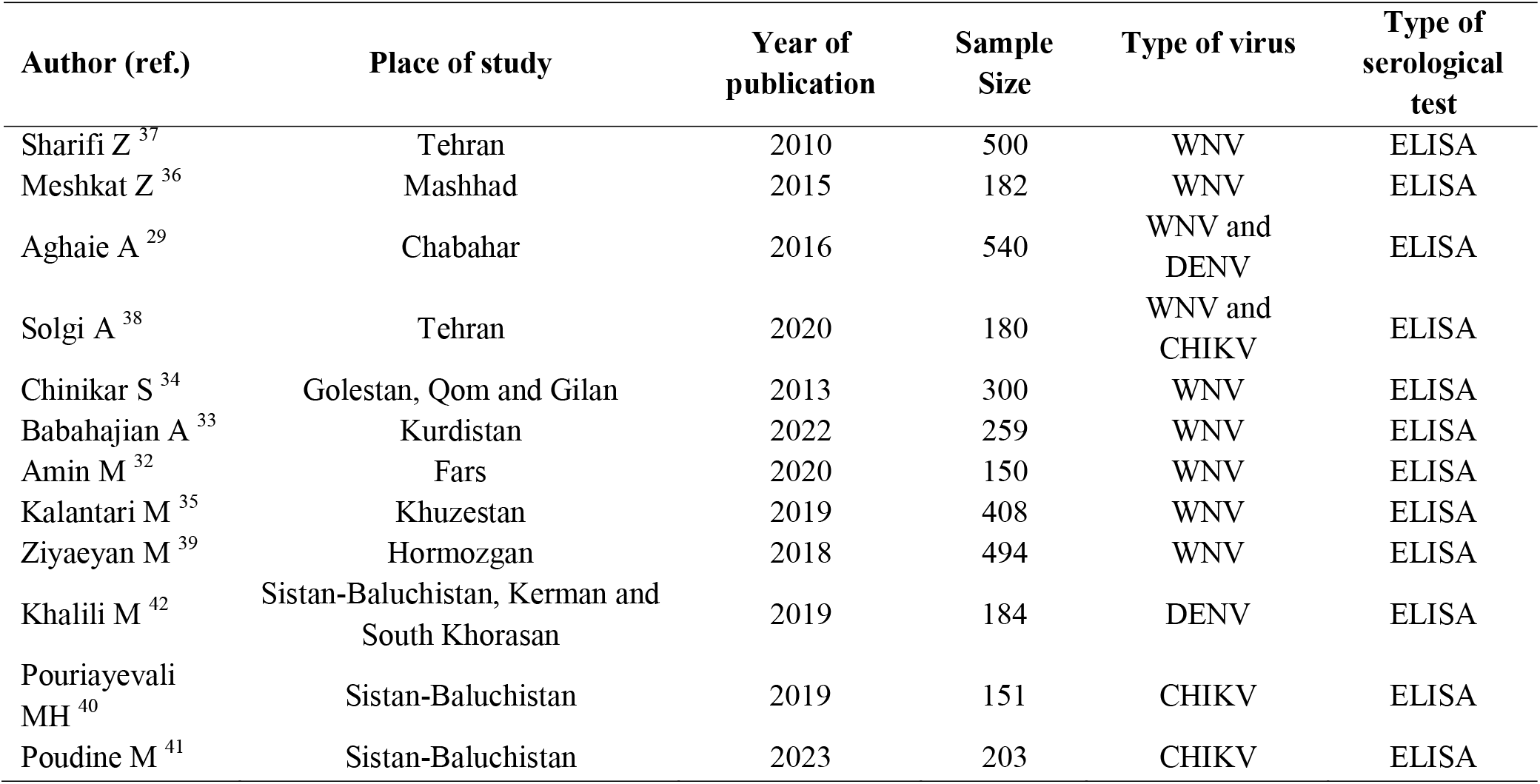
Characteristics of the articles included in the meta-analysis.

Nine articles with a sample size of 3013 people in the field of WNV serology in Iran were included in the meta-analysis process. Based on these findings, the prevalence of WNV was estimated to be 12.9% using the ELISA serology test in Iran. Among the surveyed studies, the highest prevalence of WNV positive tests was related to that conducted in Fars province with a prevalence of 27.3%, and the lowest prevalence of the test related to the studies carried out in Golestan, Gilan, and Qom provinces with a prevalence of 1.3% (Figure 2).

**Figure 2.**
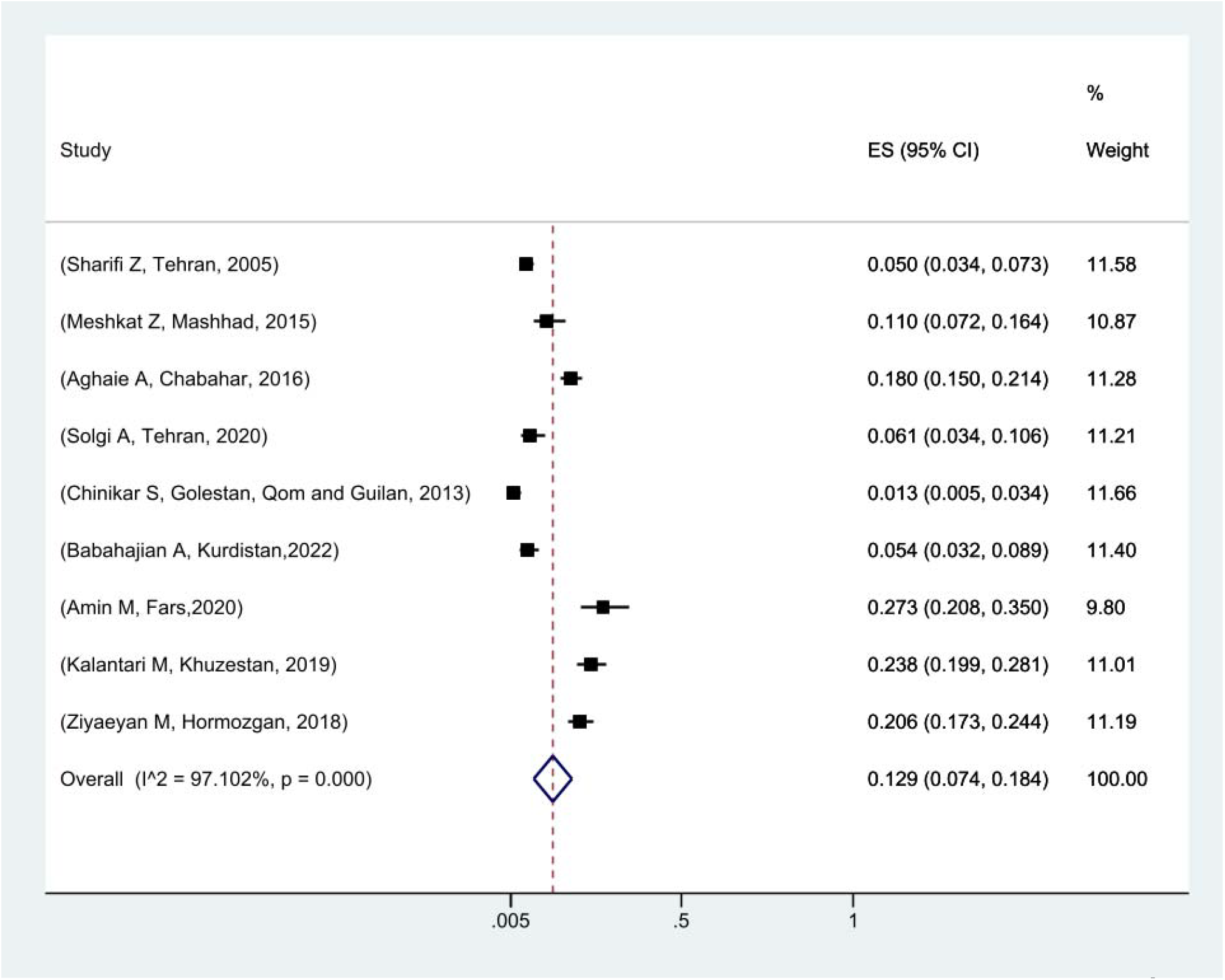
Pooled prevalence of positive ELISA test for West Nile virus in Iran based on the random effects model. The midpoint of each line segment shows the prevalence estimate, the length of the line segment indicates the 95% confidence interval in each study, and the diamond mark illustrates the pooled prevalence.

The meta-analysis of three studies performed on the prevalence of positive CHIKV assay in Iran showed that 6.2% of the studied population had a positive ELISA test in this regard (Figure 3). The prevalence of positive Dengue fever virus ELISA test in Iran was also investigated in two studies. In one survey ^42^, there was no case of positive ELISA test in the field of DENV, while the prevalence of this virus was reported at 5.6% in the other study ^43^ conducted in Sistan-Baluchistan province ^29 43^.

**Figure 3.**
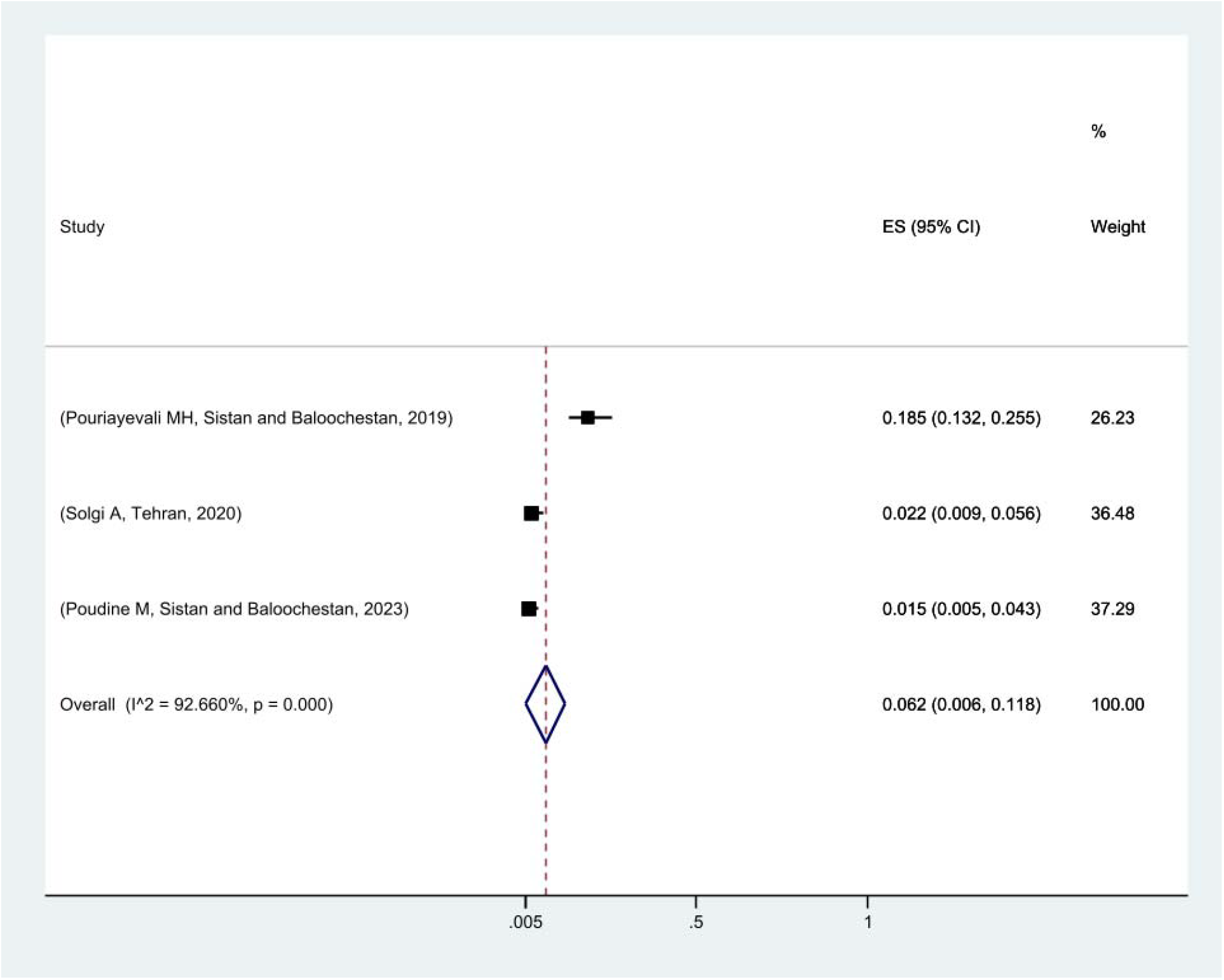
Pooled prevalence of positive ELISA test for Chikungunya virus in Iran based on random effects model. The midpoint of each line segment shows the prevalence estimate, the length of the line segment indicates the 95% confidence interval in each study, and the diamond mark illustrates the pooled prevalence.

The publication bias was evaluated using a funnel plot and Egger test. Considering the symmetry of the funnel plot and the fact that the studies with a high sample size are placed under the plot, it can be mentioned that the publication bias did not occur (*P* = 0.12) (Figure 4). Although several studies with a smaller sample size are outside the graph, it shows that to make a general consensus, it is necessary to conduct another series of studies in this field. Investigating the prevalence of positive ELISA tests based on the current sample volumes showed that with the increase in sample size, the prevalence of positive ELISA tests has also increased (Figure 5).

**Figure 4.**
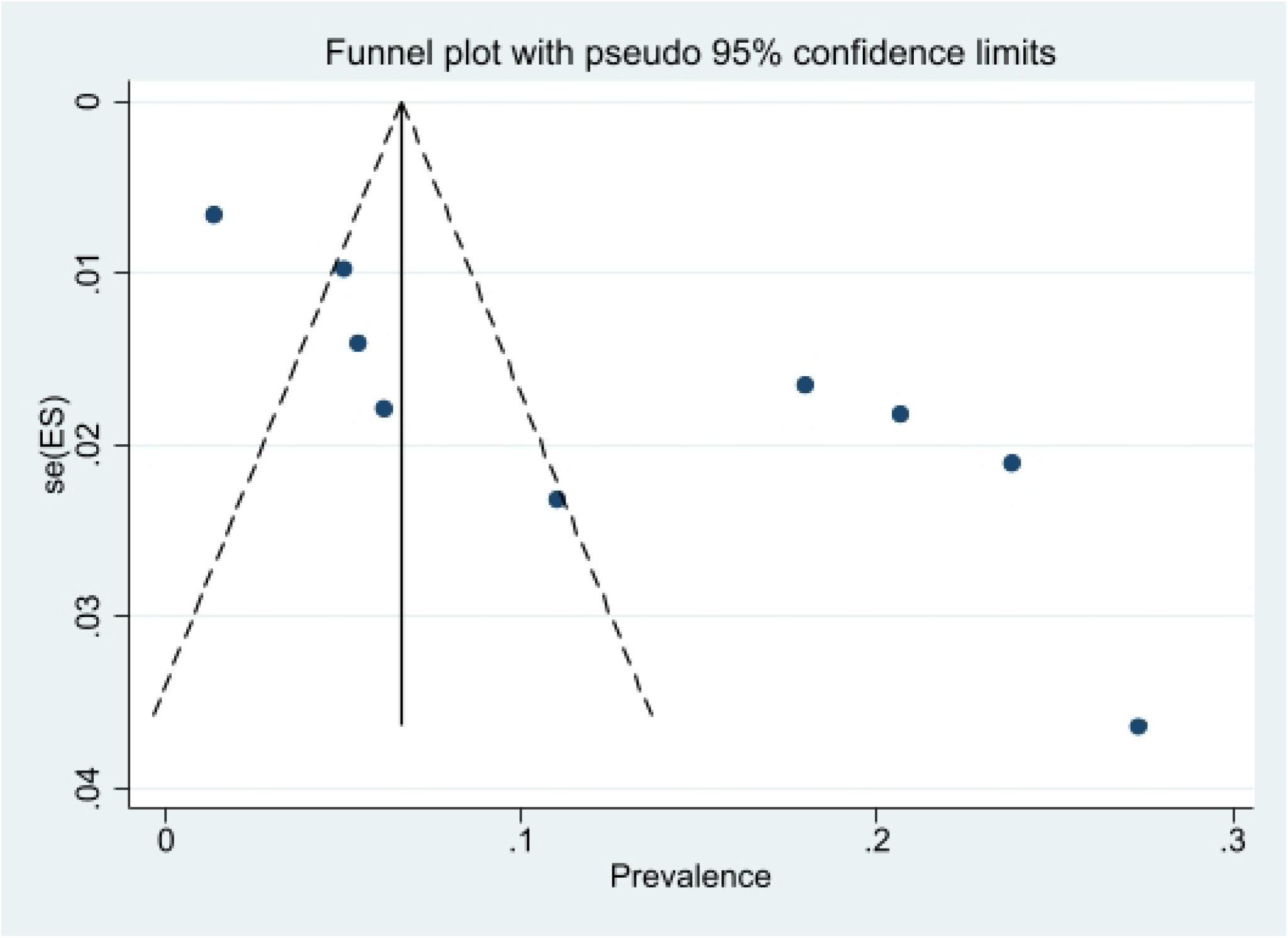
Funnel plot of the prevalence positive ELISA test in the selected studies.

**Figure 5.**
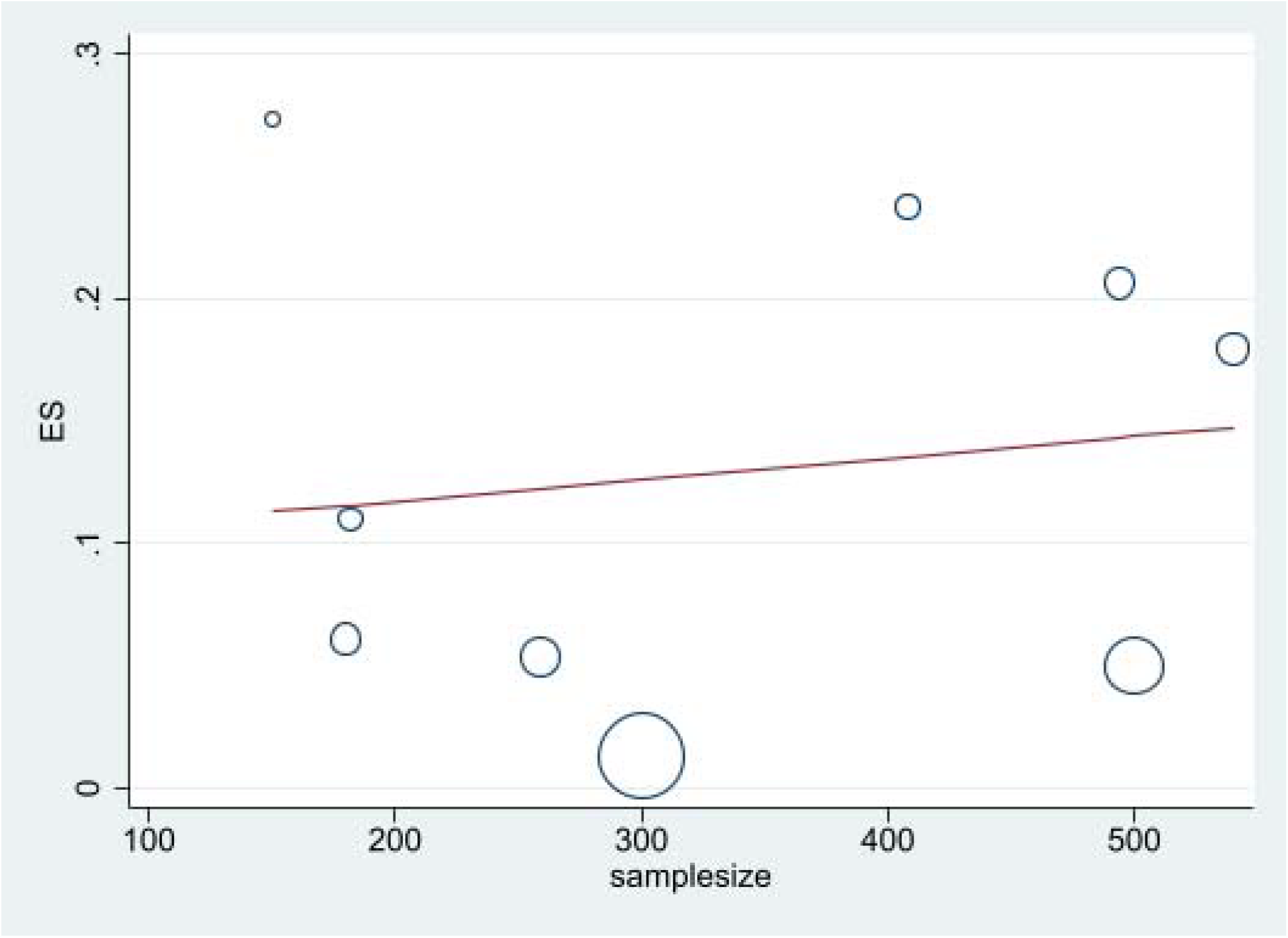
Meta-regression plot of prevalence of West Nile virus and sample size of study.

## Discussion

Based on the findings of this meta-analysis using the ELISA serology test in Iran, the WNV prevalence was estimated to be 12.9% (95%, CI: 18.4-7.4). In the past decades, WNV has spread in most regions of the world, which has affected its virulence, pathogenicity, epidemiology, and hosts. WNV has caused major epidemics with thousands of human morbidity cases and mortality in the world ^44^. This virus is endemic in the Middle East, in the countries of Pakistan, Jordan, Turkey, Iraq, Oman, Saudi Arabia, Sudan, Yemen, Egypt and Afghanistan ^45-47^. The prevalence of WNV was 26.6% in Pakistan, 30.4% in Afghanistan, and 10.4% in Qatar ^48 49^. In Iraq (11.6-15.1%), Egypt (1-61%), Jordan (8%), Iran (0-30%), Libya (2.3%), Lebanon (0.5-1%), Pakistan (0.6-65.0%), Morocco (0-18.8%), Tunisia (4.3-31.1%), and Sudan (2.2-47%) have been reported ^50^.

The WNV virus is ubiquitously endemic in Iran. Various factors, including weather conditions such as temperature and relative humidity fluctuations, can affect the activity of this virus vector, as a result, its prevalence varies in different regions of the country ^51^. In general, the prevalence of WNV in Iran is close to the endemic areas and neighboring countries of Iran. According to the results of surveys included in this meta-analysis process, the prevalence of this virus in the southern provinces was higher than in the northern and western provinces of Iran, which can be related to the increased amplification of its virus vectors in the south, and neighboring countries of Iran, which have a higher prevalence of this virus. In general, the prevalence of WNV in Iran is relatively high, and it is necessary to evaluate the circulation of this virus in Iran every year, and implement preventive measures to mitigate the spread of this virus in the country.

The results of meta-analysis on the prevalence of positive ELISA test for CHIKV showed that this virus prevalence was 6.2% (95%, CI: 0.6-11.8) in Iran. Studies have shown that the prevalence of CHIKV varies in different regions of the world. Its epidemics occur every 7-8-year period. The CHIKV is not life-threatening in contrast to DENV. The prevalence of CHIKV has been reported in Italy as 10.2% ^52^, in India as 22.3% ^53^, in Tanzania as 3.7% ^54^ and in Turkey as 0.4% ^55^. Also, the presence of CHIKV has been reported only in Saudi Arabia, Pakistan, Sudan, Yemen, Somalia, Egypt, Oman, Iraq, and Kuwait, and is known to be endemic in many parts of these regions ^8 56^. In general, it can be mentioned that this disease has a relatively high prevalence in Iran, and Iran can be considered as an endemic region for CHIKV. Although this disease causes more complications to people, and its attenuation is relatively low, knowing its prevalence is essential for control programs. Based on this, it is necessary to assess and monitor the serological prevalence of this virus annually.

With regard to Dengue fever virus (DFV), only one study which was conducted in Sistan-Baluchistan, the prevalence of this virus was 5.6% ^43^. Other studies were conducted in the form of reports of Dengue fever in Tehran (2012 and 2009) ^57 58^, and it showed that this virus was detected in different years in Iran. But apart from the SE regions of Iran, it has occurred sporadically in other regions. It should be noted that most of these cases were often reported as imported cases.

Dengue fever is recognized as a disease in developing countries of SE Asia, and most cases occur in these countries ^59^. Pakistan, Yemen, Saudi Arabia, Madagascar, and Sudan are among the countries where dengue fever is reported as endemic ^60^. In India, the prevalence of dengue fever is 23%, and in Sudan, it is 47.6% ^61 62^. These countries and SE Asian countries are often considered tourist destinations for Iranians, and many people from Iran travel to these countries every year. As a result, the possibility of contracting DFV, and transferring it inside the country increases, especially among travelers returning from Saudi Arabia following pilgrimage ^57 63^. In general, it can be mentioned that cases identified in other provinces of Iran, except Sistan-Baluchistan, can be imported cases from other countries, and often occur sporadically.

Approximately, 3.9 billion people in 129 countries are at risk of contracting DENV worldwide. Almost 70% of this global burden of DENV is related to Asia. According to World Health Organization (WHO), DENV cases have levitated more than eight times in the last two decades. Several important outbreaks of DENV have recently happened in the Eastern Mediterranean Regional Organization (EMRO) countries, including Saudi Arabia, Yemen, Oman, Sudan, and Pakistan. Alarmingly, asymptomatic humans despite low-medium level of viremia transmit DENV to vector mosquitoes ^64^. The inter-epidemic period for DENV is 3-5 years.

The main anthropophilic vector, *Aedes aegypti*, of DENV is a diurnally-active endophilic species. After seven decades, this mosquito species has re-emerged, particularly in the coastal SE region of Iran. There are also unconfirmed reports of *Aedes albopictus* presence in the same Oriental provinces of this country. Pregnant females and immunocompromised individuals, like those with diabetes, allergies, and many chronic diseases, are specially at risk of being afflicted with DENV.

## Conclusions

Based on the present systematic review and meta-analysis findings, WNV, CHIKV, and DENV have been detected in Iran using ELISA test. Considering the prevalence of WNV and CHIKV, and also the identification of these viruses in several provinces, it could be postulated that these two viruses are endemic in Iran, while DENV occurs sporadically in Iran. Based on this, to monitor and surveil the spread and outbreak of these viruses, it is recommended to screen suspected travelers and high-risk cohorts from highly-endemic neighborhood regions using the ELISA test combined with the more sophisticated molecular tools such as polymerase chain reactions (PCR).

(***N.B***. While this MS was under the submission process, three new fatal cases of DENV were reported from the SE counties of Fars and Hormozgan provinces, Iran).

## Data Availability

All data produced in the present work are contained in the manuscript.

## Ethical Approval

Not applicable. No ethical approval is applicable since it is a review article.

## Data Availability and Materials

The data that support the findings of this study are available from the corresponding author upon reasonable request

## Acknowledgments

The authors wish to convey their appreciation to the Vice-Chancellor for Research and Technology at Shiraz University of Medical Sciences (SUMS) for provision of software support and other logistics. This meta-analysis report was part of a self-initiative attempt to screen and elucidate the current status of three highly-important clinical arboviruses in Iran, forming a basis to the part fulfilment of a PhD thesis by E.A. under the supervisorship of M.D.M-F. in the Dept. of Biology and Control of Disease Vectors, School of Health, Shiraz, Iran.

